# SARS-CoV-2 in first trimester pregnancy – does it affect the fetus?

**DOI:** 10.1101/2020.06.08.20125195

**Authors:** Nina la Cour Freiesleben, Pia Egerup, Kathrine Vauvert Römmelmayer Hviid, Elin Rosenbek Severinsen, Astrid Marie Kolte, David Westergaard, Line Fich Olsen, Lisbeth Prætorius, Anne Zedeler, Ann-Marie Hellerung Christiansen, Josefine Reinhardt Nielsen, Didi Bang, Sine Berntsen, Joaquim Ollé-López, Andreas Ingham, Judith Bello-Rodríguez, Ditte Marie Storm, Jeppe Ethelberg-Findsen, Eva R Hoffmann, Charlotte Wilken-Jensen, Finn Stener Jørgensen, Henrik Westh, Henrik Løvendahl Jørgensen, Henriette Svarre Nielsen

## Abstract

**Background:** Several viral infections are known to be harmful to the fetus in the first trimester of pregnancy and can cause increased nuchal translucency thickness and pregnancy loss. Currently, no evidence exists regarding possible effects of SARS-CoV-2 in first trimester pregnancies.

**Methods:** Cohort 1 included pregnant women with a double test taken between Feb. 17 and Apr. 23, 2020, during the SARS-CoV-2 epidemic peak in Denmark. The double test was taken as part of the first trimester risk assessment. Cohort 2 included women with a first trimester pregnancy loss before double test. Serum from the double test or from a blood sample, in case of pregnancy loss, was analyzed for SARS-CoV-2 antibodies. The results were correlated to the nuchal translucency thickness and the number of pregnancy losses.

**Results:** In total, 1,019 pregnant women with double test and 36 women with pregnancy loss participated in the study. Thirty (2.9%) women had SARS-CoV-2 antibodies in the serum from the double test. All women with pregnancy loss prior to the double test were negative for SARS-CoV-2 antibodies. There were no significant differences in nuchal translucency thickness for women testing positive (n=14) versus negative (p=0.20) or grey zone (n=16) versus negative (p=0.28). In total, 54 women experienced a pregnancy loss of whom two had grey zone or positive SARS-CoV-2 antibodies.

**Conclusion:** Maternal SARS-CoV-2 infection did not seem harmful in first trimester pregnancies. Infection had no effect on the nuchal translucency thickness and women with SARS-CoV-2 antibodies were not overrepresented among women with pregnancy loss.

## Introduction

The first case of Coronavirus disease 2019 (Covid-19) was reported in Wuhan, China, in December 2019 and within a few months it developed into a worldwide pandemic currently impacting 188 countries.^1^ Covid-19 is caused by severe acute respiratory syndrome coronavirus-2 (SARS-CoV-2). As of June 5, 2020, more than 6.5 million people worldwide were infected resulting in 387,155 deaths.^2^

Pregnant women are more vulnerable to viral infections and therefore represent a potential risk group for severe outcomes in relation to viral infections.^3^ Especially, they have an increased risk of severe pneumonia following infections with respiratory pathogens.^4^ The increased susceptibility during first trimester pregnancy may be due to a pro-inflammatory state.^4^

For pregnant women, previous coronavirus epidemics such as middle east respiratory syndrome (MERS) and severe acute respiratory syndrome (SARS) have been associated with increased maternal morbidity, mortality and adverse obstetric outcomes.^5^ Only a few documented cases of SARS in pregnant women have been reported. A case study from Hong Kong of seven first trimester cases showed a pregnancy loss rate of 57% in women infected with SARS.^6^ Only 11 confirmed cases with MERS infection during pregnancy have been documented worldwide showing a maternal- and infant fatality rate of 27%.^5,7^ Thus, there is a general paucity of data on which to base public health policies for pregnant women and risks associated with SARS-CoV-2 infection.

Vertical maternal-fetal transmission with serious fetal consequences can occur in relation to maternal infection with TORCH agents (Toxoplasma, Other, Rubella, Cytomegalovirus, Herpes) and Zika virus.^3,8,9^ As the fetal organs develop during the first trimester of pregnancy, maternal infections at this stage may be more severe compared to later gestational ages.^3,8^ Likewise, Parvovirus B19 infection during pregnancy, even in asymptomatic women, is associated with an increased nuchal translucency thickness^10^ and can be harmful for the fetus. Vertical transmission in relation to SARS and MERS has not yet been documented.^9^

Evidence concerning Covid-19 in pregnancy is still limited and serological testing for SARS-CoV-2 antibodies has only been reported in one study of six pregnant women in the third trimester of pregnancy.^11^ To our knowledge, no studies have yet investigated Covid-19 in first trimester pregnancies.

In this study, we used serological testing to identify women with SARS-CoV-2 infection in early pregnancy to investigate the impact of Covid-19 on the fetus in first trimester pregnancies focusing on signs of fetal infection (nuchal translucency scan) and pregnancy loss.

## Methods

All pregnant women in Denmark are offered a combined first trimester risk assessment (performed at gestational age 11-14) as part of the public antenatal and obstetric health care service, free of charge. More than 90% of the women accept.^12^ The risk assessment includes a double test (blood sample for pregnancy-associated plasma protein A (PAPP-A) and free beta human chorionic gonadotropin (β-hCG)) and a nuchal translucency measurement with ultrasonography. The excess serum from the double test is stored at minus 80 degrees Celsius at the hospital.

### Participants

The Department of Obstetrics and Gynecology at Copenhagen University Hospital Hvidovre is the largest obstetric department in Denmark serving approximately 12% of pregnant women and births in Denmark. All pregnant women, who had a double test performed at Hvidovre Hospital from February 17, 2020 to April 23, 2020 were invited to participate in the study. The women were contacted electronically with written information about the study. If they agreed to participate an informed consent form was signed and the women were included in the study (Cohort 1). We included women who consented up until May 28.

From April 14 to May 21, 2020, women referred with a pregnancy loss before the time of the double test, were also invited to participate. If a woman with a pregnancy loss wanted to participate, a blood sample was drawn, and baseline characteristics were collected by cross-referencing medical files (Cohort 2).

A short questionnaire concerning symptoms of Covid-19 during the pregnancy, smoking habits, body mass index (BMI), influenza vaccination in 2019/2020 and comorbidity was completed by all participating women.

### Antibody analysis

The stored excess serum from the double tests and the blood samples from women with pregnancy loss, 30 µl serum from each sample, were analyzed for antibodies (IgM and IgG) against SARS-CoV-2.

The serological antibody test for SARS-CoV-2 is an indicator of previous SARS-CoV-2 infection.^13^ Antibodies may be present from day four following the first symptoms^14^ and the median seroconversion time is day-12 for IgM and day-14 for IgG.^15^

Samples were analyzed using YHLO’s iFlash 1800 and SARS-CoV-2 IgM/IgG kits. In accordance with the recommendations of the manufacturer, IgM and IgG antibody values ≤8 AU/mL were defined as negative results and values ≥12 AU/mL were defined as positive results. Values >8.0 and <12.0 AU/mL were considered “grey zone” results.^16^

### Statistical analysis

Data and figures were analyzed and produced using R, an open source statistical software (the R foundation, www.r-project.org). Two-way comparisons of nuchal translucency thickness, free β-hCG, and PAPP-A between women with and without SARS-CoV-2 antibodies were performed using the Wilcoxon Rank Sum test. Multivariable modelling of the effect of Covid-19 infection in first trimester pregnancy on nuchal translucency thickness, free β-hCG and PAPP-A was performed using an ordinal regression model, taking maternal age and gestational age into account. Differences in reported Covid-19 symptom frequency were analyzed using Fishers exact test. A p-value < 0.05 was considered statistically significant.

### Ethics

The study was approved by Knowledge Centre on Data Protection Compliance, The Capital Region of Denmark (P-2020-255) and by the Scientific Ethics Committee of the Capital Region of Denmark (journal number H-20022647). All participants in the study provided written informed consent.

## Results

A total of 1,356 double tests were performed from 1,356 pregnant women during the study period. Of the 1,356 women, 1,019 (75.1%) provided informed consent to participate (Cohort 1). Additionally, 36 women with an early pregnancy loss prior to the time of the double test were included (Cohort 2). The overview of the study is illustrated in Figure 1.

**Figure 1.**
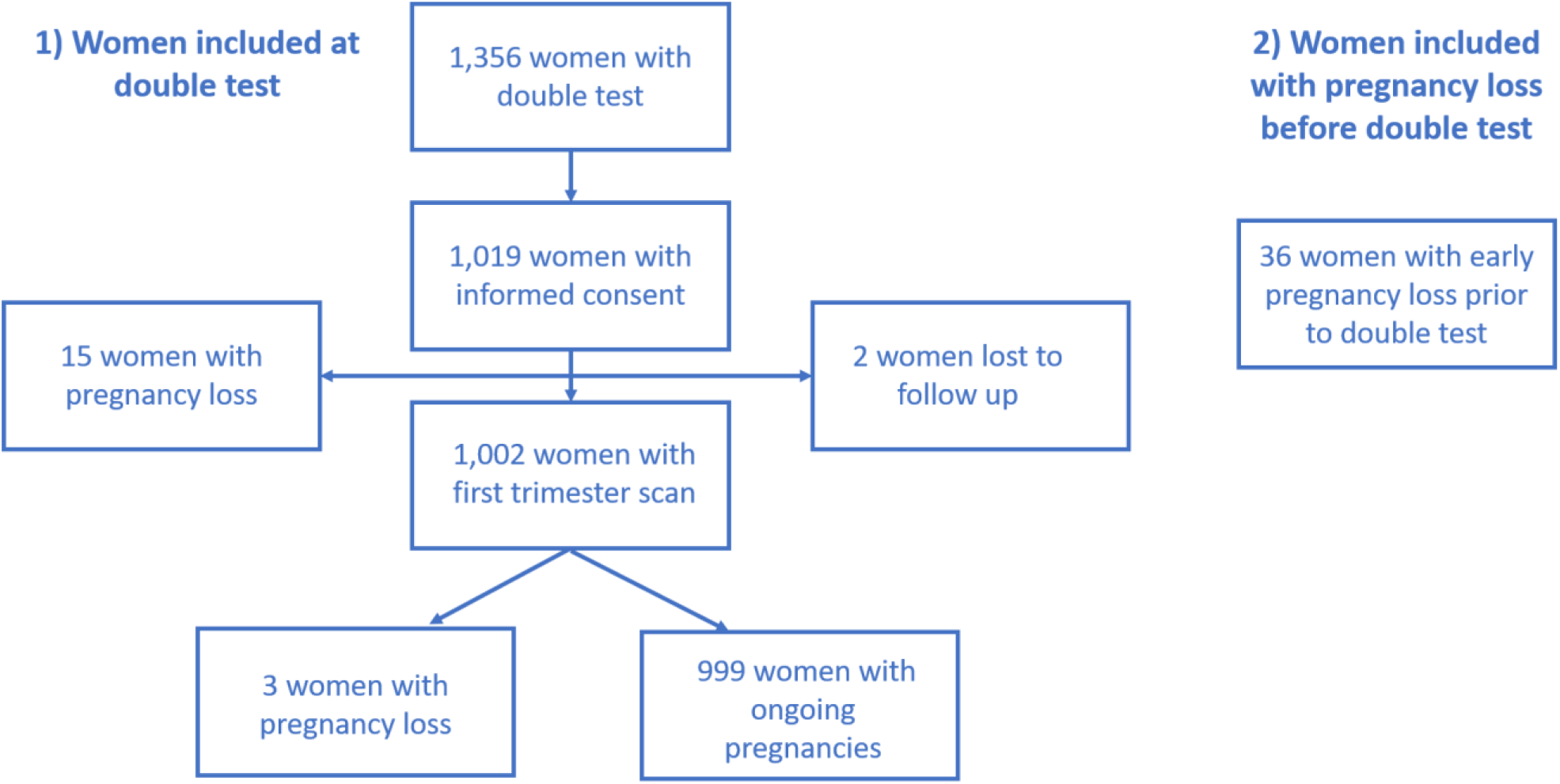
Flowchart of the two cohorts

The median gestational age was 11 weeks and 0 days (11+0) at the double test and 13+0 at first trimester scan. The median gestational age among the 36 women with early pregnancy loss was 8+1. The characteristics of the two cohorts are presented in Table 1.

**Table 1.** Characteristics of the included women in the two cohorts

|  | Women included<br>after double test<br>n = 1,019<br>(Cohort 1) | Women with pregnancy loss before<br>double test<br>n= 36<br>(Cohort 2) |
| --- | --- | --- |
| **Age, mean (SD)** | 31.71 (4.52) | 32.96 (5.22) |
| **BMI, mean (SD)** | 23.92 (4.65) | 25.33 (5.51) |
| **Gestational week, median** |  |  |
| - At blood sample for double test | 11+0 |  |
| - At first trimester scan (nuchal translucency) | 13+0 |  |
| - At pregnancy loss |  | 8+1 |
| **Smoking, n (%)** |  |  |
| - Yes | 31 (3.0) | 3 (8.3) |
| - No | 953 (93.5) | 31 (86.1) |
| - Unknown | 35 (3.4) | 2 (5.6) |
| **Asthma, n (%)** |  |  |
| - Yes | 56 (5.5) | 4 (11.1) |
| - No | 942 (92.4) | 31 (86.1) |
| - Unknown | 21 (2.1) | 1 (2.8) |
| **Influenza vaccination 2019/2020, n (%)** |  |  |
| - Yes | 99 (9.7) | 5 (13.9) |
| - No | 900 (88.3) | 23 (63.9) |
| - Unknown | 20 (2.0) | 8 (22.2) |
| **SARS-CoV-2 antibodies, n (%)** |  |  |
| - Negative | 989 (97.1) | 36 (100) |
| - Grey zone | 16 (1.6) | 0 |
| - Positive | 14 (1.4) | 0 |

The total number of women with SARS-CoV-2 antibodies (grey zone and positive IgM and/or IgG values) in Cohort 1 was 30 (2.9%). 14 women (1.4%) had a positive antibody level and 16 women (1.6%) had a grey zone SARS-CoV-2 antibody level. Two women were IgM-only positive, one woman was IgG and IgM positive, and one was IgG positive and IgM grey zone. Of the remaining 26 women, 10 were IgG-only positive, 10 had grey zone IgG values, and 6 patients had grey zone IgM values. None of the 36 women from Cohort 2 had positive or grey zone SARS-CoV-2 antibody levels.

We subsequently assessed the association of SARS-CoV-2 antibody levels in Cohort 1 with the first trimester scan (nuchal translucency thickness) and the double test. Women, where the fetus was found to have a chromosome anomaly (trisomy), were excluded from the analysis comparing nuchal translucency thickness and SARS-CoV-2 antibody level. The median nuchal translucency thickness, free β-hCG, and PAPP-A levels were not significantly different between women with negative versus positive levels of SARS-CoV-2 antibodies (Table 2). Additionally, there was no significant difference in nuchal translucency thickness for women with grey zone (n=16) versus negative antibodies (p=0.28). Also, after accounting for maternal age and gestational week, positive antibodies (p=0.47) or grey zone antibodies (p=0.21) did not affect nuchal translucency thickness.

**Table 2.**
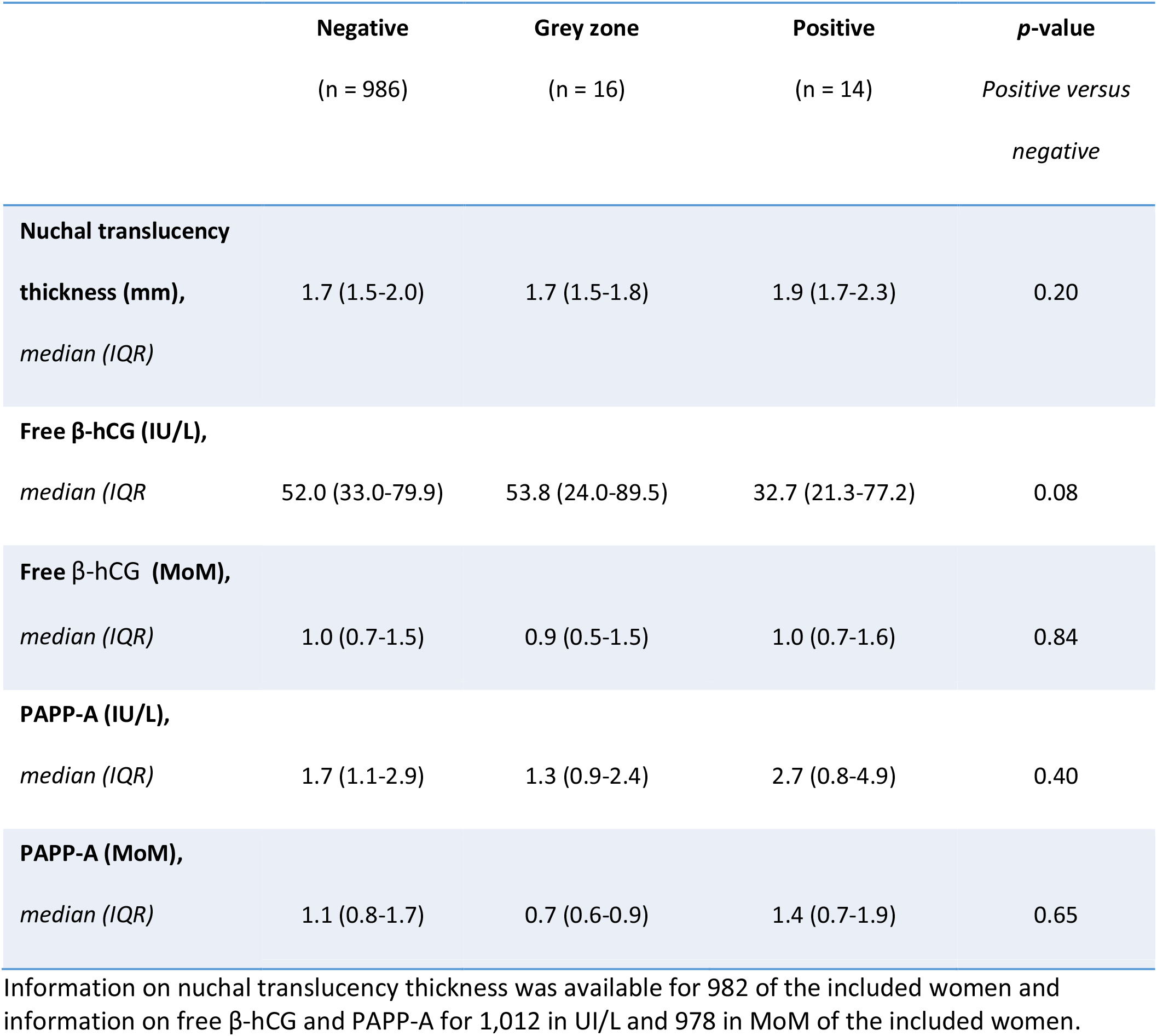
Primary outcomes for cohort 1, which was included after the double test. Outcomes are stratified according to the levels of SARS-CoV-2 antibodies (IgM and IgG).

Table 3 displays pregnancy status for all 1,055 pregnancies (1,019 in Cohort 1 and 36 in Cohort 2) after the first trimester and according to SARS-CoV-2 antibodies. 36 women had a pregnancy loss before the double test (Cohort 2), 15 women had a pregnancy loss between the double test and the nuchal translucency scan, and three women were diagnosed with a missed abortion at the nuchal translucency scan (Figure 1). This totals 54 first trimester pregnancy losses. Two women with grey zone or positive SARS-CoV-2 antibody levels had a pregnancy loss and 27 women with ongoing pregnancies had SARS-CoV-2 grey zone or positive antibody levels. Two women were lost to follow up after the double test.

**Table 3.** Pregnancy status after the first trimester according to SARS-CoV-2 antibody levels. The table includes both Cohort 1 and 2.

|  | **Negative**<br>(n = 1,024) | **Grey zone**<br>(n = 16) | **Positive**<br>(n = 13) |
| --- | --- | --- | --- |
| **Ongoing pregnancy, *n*** | 972 | 15 | 12 |
| **Pregnancy loss, *n*** | 52 | 1 | 1 |

One woman with a positive test result and one woman with a negative test result were lost to follow up after double test.

Figure 2 illustrates Covid-19 symptoms reported by pregnant women with negative, grey zone or positive SARS-CoV-2 antibody levels. Significantly more women with grey zone antibody levels reported symptoms compared to women with negative antibody levels (OR=4.94, 95%CI 1.61-16.73). Women with positive antibody levels did not report more symptoms compared to women with negative antibody levels (OR=2.23, 95%CI 0.63-7.40).

**Figure 2.**
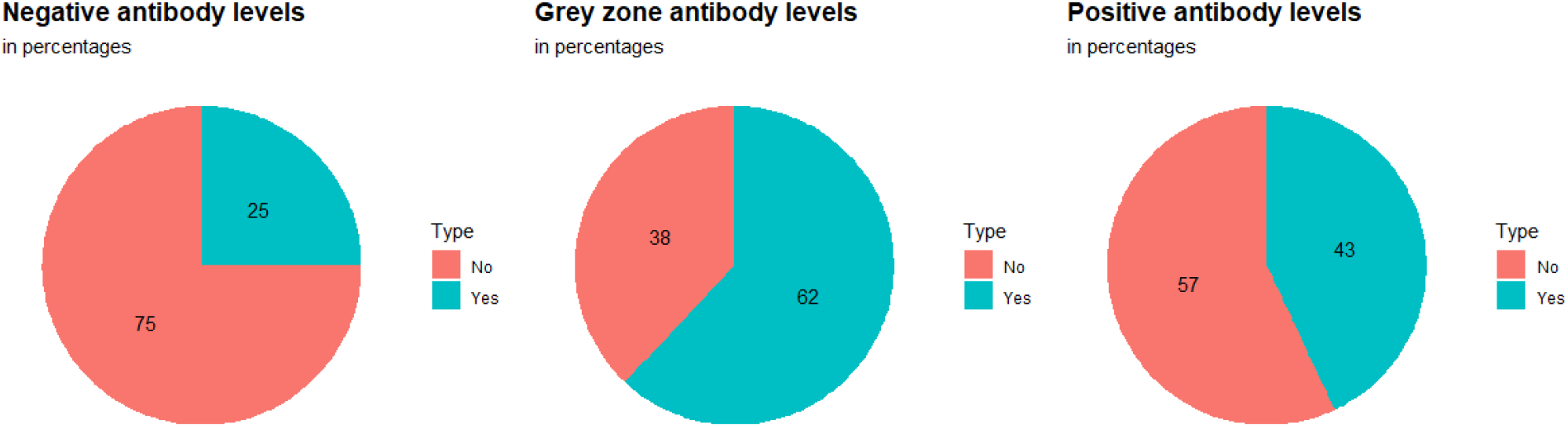
Incidence of self-reported Covid-19 symptoms for women with positive, grey zone or negative SARS-CoV-2 antibodies (IgM and IgG).

The cumulative frequency of pregnant women included after the double test (Cohort 1) and with a grey zone or positive SARS-CoV-2 antibody level during the study period is displayed in Supplementary Figure 1.

## Discussion

We found that pregnant women with SARS-CoV-2 infection in the first trimester did not have a significantly different nuchal translucency thickness measured at their first trimester scan. Of the 36 women with early pregnancy loss, before the double test was taken, none had SARS-CoV-2 antibodies. We included Cohort 2 to minimize the risk of bias that might potentially be caused by pregnancy loss early in the first trimester in infected women. Only two women with pregnancy losses after the double test had SARS-CoV-2 antibodies. None of the 30 women with SARS-CoV-2 antibodies had been hospitalized for Covid-19.

The first case of Covid-19 in Denmark was confirmed Feb. 27, 2020. At the beginning of the epidemic in Denmark, it was only individuals requiring hospitalization who were tested for SARS-CoV-2 with a respiratory specimen. Citizens suspected of Covid-19 but not requiring admission were asked to remain at home and self-quarantine and were not tested. Only about half of the women with SARS-CoV-2 antibodies in our study reported symptoms of Covid-19 in pregnancy. Therefore, serological testing in appointed risk groups, such as pregnant women, is a valuable tool to identify previous infections and to evaluate whether infection in pregnant women requires additional vigilance during the pregnancy. Our findings suggest that pregnant women in their first trimester are not at increased risk of severe Covid-19 disease. This is similar to what has been reported for pregnant women in their third trimester in Wuhan, China.^17^

In general, the study population had normal BMI and the vast majority were non-smokers. Our results and conclusion may therefore not apply directly to populations with higher BMI, higher frequency of smoking and associated higher frequency of lifestyle diseases. People with lifestyle diseases and individuals who smoke are at higher risk of developing more severe Covid-19 disease if infected^18–20^. Our study does not rule out the possibility that more severe Covid-19 disease might lead to a higher risk of adverse outcomes for the developing fetus.

The frequency of participants with a positive or grey zone SARS-CoV-2 antibody test was relatively low and steady over the study period (Supplementary Fig. 1). This is most likely a result of the extended measures implemented by the Danish government at an early stage of the epidemic to limit the transmission of the virus. Measures included closing the national borders, banning of group gatherings of more than ten people, closure of all educational facilities, implementing work-from-home measures for all non-critical government and state employees, and recommending that private employees also work from home where feasible. However, in addition to the general societal changes, the relatively low occurrence of SARS-CoV-2 antibodies among participants could also be due to pregnant women taking additional precautionary measures such as self-quarantine and limiting social contacts even before the implementation of official governmental restrictions. As per May 23, 2020, it was estimated that the seroprevalence of people with SARS-CoV-2 antibodies in the Danish population was 1.1% (95%CI 0.5–1.8).^21^ This frequency fits well with the findings of this study. However, it should be noted that at the time of writing, only 41.2% of 2,600 randomly selected citizens had been recruited and sampled.^21^

There is still uncertainty concerning the accuracy of the various tests for SARS-CoV-2 antibodies.^22^ One study from China used a sandwich enzyme linked immunosorbent assay^14^ and found the sensitivity and specificity to be 77.3%/100% for IgM and 83.3%/95% for IgG, respectively. We used iFlash 1800 with its IgM/IgG kit, which has previously shown highly accurate results.^13^ However, it is possible that even more accurate assays will be developed soon. As reviewed by Infantino *et al*., more studies are needed to validate the serological assays, especially for use as screening tools for asymptomatic individuals.^22^ Our study also includes a risk of bias due to the relatively low prevalence of positive samples. This may increase the risk of false positive test results and therefore, the “true” prevalence could be lower than estimated.^23^

Another potential limitation of the study is that not all invited women participated in the study. By the end of May 28, 2020, a total of 337 women had not responded to our study invitation. It could potentially introduce selection bias if the none-respondents were different in terms of rates of positive SARS-CoV-2 antibodies and pregnancy loss. To mitigate the risk of such bias, we included Cohort 2. The antibody analyses from these women’s blood samples, which were drawn at the day of pregnancy loss, did not show a high frequency of SARS-CoV-2 antibodies. This provides some certainty that we did not overlook a potential effect of SARS-CoV-2 at the earlier stage of pregnancy before the double test is taken.

## Conclusion

This is the first study investigating the impact of SARS-CoV-2 on first trimester pregnancies. We did not find a higher median nuchal translucency thickness at the first trimester scan among women with positive SARS-CoV-2 IgM or IgG antibodies than among women without SARS-CoV-2 antibodies. Women with SARS-CoV-2 antibodies were not overrepresented among women with pregnancy loss before the double test. Serological studies investigating the impact of SARS-CoV-2 on later stages of pregnancy are needed to develop clinical guidelines and recommendations for any possible restrictions for pregnant women in relation to SARS-CoV-2.

## Data Availability

All data referred to in the manuscript are included in the approved database for this study

## Acknowledgements

We thank all the laboratory technicians who helped to collect and process the blood samples for the study.

